# Differences in bacterial colonization and mucosal responses between high and low SES children in Indonesia

**DOI:** 10.1101/2021.11.30.21267078

**Authors:** Marloes M.A.R. van Dorst, Shohreh Azimi, Sitti Wahyuni, Aldian I. Amaruddin, Erliyani Sartono, Linda J. Wammes, Maria Yazdanbakhsh, Simon P. Jochems

**Author notes:** correspondence to **Corresponding author information** Dr. Simon P. Jochems, Department of Parasitology, Leiden University Medical Center, Albinusdreef 2, 2333 ZA Leiden, The Netherlands. contributed equally (Marloes M.A.R. van Dorst, Shohreh Azimi).

## Abstract

**BACKGROUND:** Nasopharyngeal carriage of pathogenic bacteria precedes invasive disease and higher rates are found in low socioeconomic-status (SES) settings. Local immune responses are important for controlling colonization, but whether SES affects these responses is currently unknown.

**OBJECTIVE:** Examining bacterial colonization and cytokine response in nasal mucosa of children from high and low SES.

**METHODS:** Twenty-five cytokines were measured in nasal fluid. qPCR was performed to determine carriage and density of *Haemophilus influenzae* (H. influenzae), *Streptococcus pneumoniae* (S. pneumoniae), *Moraxella catarrhalis* (M. catarrhalis) and *Staphylococcus aureus* (S. aureus).

**RESULTS:** The densities of H. influenzae and S. pneumoniae were increased in low compared to the high SES (*p*=0.006, *p*=0.026), with respectively 6 and 67 times higher median densities. Densities of H. influenzae and S. pneumoniae were positively associated with levels of IL-1beta (*p*=0.002, *p*=0.008) and IL-6 (*p*<0.001, *p*=0.006). After correcting for bacterial density, IL-6 levels were increased in colonized children from high compared to low SES for both H. influenzae and S. pneumoniae (both *p*=0.039).

**CONCLUSION:** Increased density of H. influenzae and S. pneumoniae was observed in low SES children, while IL-6 levels associated with colonization were reduced in these children, indicating that immune responses to bacterial colonization were altered by SES.

## Introduction

Lower respiratory tract infections (LRTIs) remain the leading cause of mortality amongst children under five, accounting for 2.4 million deaths in 2016 most of them in low- and middle income countries (LMICs) (1, 2).

A number infectious agents can establish LRTI, prevalent bacterial causes include *Streptococcus pneumoniae* (S. pneumoniae), *Haemophilus influenzae* (H. influenzae), *Moraxella catarrhalis* (M. catarrhalis) and *Staphylococcus aureus* (S. aureus). Nasopharyngeal carriage of these bacteria is common in infants and colonization by these bacteria is considered an essential first step in development of disease (3-5). Moreover, nasopharyngeal carriage and increased densities of S. pneumoniae, H. influenzae and M. catarrhalis have been associated with pneumonia in Tanzanian children and increased H. influenzae densities were found among children with (very) severe pneumonia in seven LMICs (6, 7).

Control of nasopharyngeal bacterial colonization is mediated by local immune responses and impaired innate cytokine responses have been associated with persistent bacterial colonization. A human inoculation study showed that clearance of nasal S. aureus requires an upregulation of chemokines, growth factors and inflammatory cytokines and that a low IL-1RA/IL-1beta ratio associates with S. aureus persistence (8). Furthermore, an increased prevalence of nasopharyngeal colonization with M. catarrhalis was observed infants with anemia, while the pro-inflammatory cytokine responses were reduced upon in vitro stimulation (9). Finally, murine models show that IL-1 signaling is essential for clearance of S. pneumoniae and suggest that reduced IL-1 responses might be permissive for persistent colonization during infancy (10).

Increased carriage rates have been observed in various settings and are associated with lower socio-economic status (SES). High nasopharyngeal bacterial colonization rates have been observed in low-income countries (11), rural areas (12) and indigenous populations (4). Furthermore, a study in Israel showed that SES rather than ethnicity drove these differences in pneumococcal carriage (13). Finally, socio-economic factors were identified as risk factors for carriage of potential pathogenic bacteria in Indonesian children (14).

Since increased bacterial colonization has been observed in low SES settings and local immune responses play an important role in the control of the bacterial colonization, reduced mucosal immune responses in low SES populations could play a role. Studies comparing immune responses between socio-economic settings are scarce and often focus on the systemic immune system. To our knowledge, no studies have compared the immune responses to bacterial colonization between different socio-economic settings. Moreover, most studies that found an association between SES and colonization have been performed in a heterogenous population. Therefore, this study aimed to examine the cytokine response to nasopharyngeal bacterial colonization of children from high and low SES schools in a single urban area.

## Methods

### Data collection

The data was obtained as part of a larger study into the effects of SES in school-aged children in Makassar, Indonesia (15). This cross-sectional study included children attending two primary schools in Makassar, the capital of South Sulawesi. These schools were selected based on the socio-economic background of the children attending the school. One of the schools is attended by children from High SES families, a privately funded school, whereas the other school is fully government funded and predominantly attended by low SES children. Both schools are located in the center of Makassar, 2km apart.

Samples were collected during September-October 2019. Informed written consent was obtained from primary caregivers. All children with written consent were included in the study, except for children using antihistamines/corticosteroids. Ethical approval was obtained from the Health Research Ethical Committee, Faculty of Medicine, Hasanuddin University (No:703/H4.6.4.5.31/PP36/2019). Socio-demographic information was obtained from all participants via questionnaires.

### Nasosorption samples preparation

Nasosorption samples were collected by inserting an absorptive matrix strip (Nasosorption™, Hunt Developments) into one nostril and pushed against the nasal lining for 30-60 seconds (16). These strips were then placed on ice until storage at −80°C within 8 hours after collection. The nasosorption samples were eluted by adding 100µL sterilized PBS with 1%BSA+0.05%Triton-X100 to the filter and spun down at 3600xg for 10 minutes at 4°C. The supernatant was transferred to a new tube and centrifuged at 16000xg, 4°C, 10 minutes. The supernatant was moved to a new tube for cytokine analysis and the pellet was stored at −20°C until DNA extraction.

### Cytokine concentration measurement

Of the supernatant, 12 µl was used to measure the concentration of 25 cytokines, by the Human Cytokine 25-plex ProcartaPlex Panel (Invitrogen, ThermoFisher No:EXP250-12166-901) according to manufacturer’s instructions, but additional washing steps and alcohol flushes were performed to account for the muckiness. The samples were measured by the Luminex200 device at normal RP1 target and concentrations were obtained by using the xPonent3.1 software in pg/mL. Children for who >90% of the cytokines could not be measured in their nasosorption sample were excluded from further analysis. This was the case for 15% of the children, 6 high and 9 low SES children.

### Generation of standard curves

The DNA concentrations of S. aureus (ATCC43300), H. influenzae (ATCC49766), S. pneumoniae 6305 and clinical isolate of M. catarrhalis were determined spectrophotometrically with the NanoDrop1000 (Thermo scientific). The copy numbers of template were calculated using the genome length computed with URI Genomics & Sequencing Center (http://cels.uri.edu/gsc/cndna.html). A standard curve was generated of 10-fold serial dilution scheme ranging from 10^7^-10^1^ copy/mL

### Bacterial colonization measurement

To determine nasal bacterial colonization, qPCR was performed on the bacterial pellet. DNA extraction was performed using magnetic beads, as previously published (17). To determine the colonization of S. pneumoniae and H. influenzae, qPCR was performed using the LytA and IgA1 gene respectively, as previously described (17, 18). qPCR for the copB gene was performed to determine colonization of M. catarrhalis and for S. aureus the nuc gene was used as previously described (6). Using the standard curves, the genome length and Avogadro’s number, the number of genome copies/µL was determined in the samples per bacterium. Details regarding the primers and probes utilized in this study can be found in supplementary ***(TableS1)***.

### Statistical analysis

The standardized z-scores of body mass index (z-BMI) were determined according to the WHO guidelines (19). To obtain approximately normally distributed data, cytokine concentrations and bacterial loads were log_10_-transformed. A Student’s t-test was performed for continuous data and for binary and categorical data Pearson’s Chi-square was used. Pearson’s correlation was used to examine the correlation of two continuous variables. To investigate the effect of SES on the bacterial loads and to estimate the effect of bacterial density and SES on cytokine levels, a regression model was performed. For all regression models in this study age, sex and z-BMI were considered *a priori* confounders and adjustment was performed accordingly. To identify the main drivers of the correlation between bacterial density and cytokine concentrations, a canonical correlation analysis was performed by using Wilk’s Lambdas including the density of the four bacteria and the concentrations of all widely measurable cytokines. The two bacteria and two cytokines that showed the largest coefficient in the canonical correlation analysis were selected for further analysis. P-values smaller than 0.05 were considered statistically significant. Data was analyzed and visualized using RStudio and R software.

## Results

### Bacterial density of H. influenzae and S. pneumoniae increased in low SES

A total of 98 school-aged children were included, 50 children attended the low SES school and 48 children the high SES school ***(TableS2)***. There was no significant difference in age or sex, but z-BMI and parental income were between lower in low SES compared to high SES children. The carriage rate was highest for M. catarrhalis, lowest for S. aureus and a combination of two or three bacteria was most common. Neither the carriage rates of the bacteria nor the number of co-colonizing bacteria differed between high and low SES.

Although the carriage rates and combinations did not differ between high and low SES, differences in the densities of bacteria were observed ***(Figure1)***. The median bacterial load of H. influenzae was 59,075 copies per nasosorption sample in low and 9,804 in high SES, indicating a 6-times higher median density. For S. pneumoniae, the median bacterial load was 97,422 in the low and 1,446 in the high SES, corresponding to a 67-times higher median density. No significant differences were observed in the bacterial loads of M. catarrhalis and S. aureus between high and low SES. To further examine the relation between the bacterial load and SES, a multivariate regression analysis was performed adjusted for a priori confounders. These results showed that SES significantly affected the H. influenzae (β_*SES*_=-1.19, *p*=0.006) and S. pneumoniae density (β_*SES*_ =-1.25, *p*=0.026).

**Figure 1:**
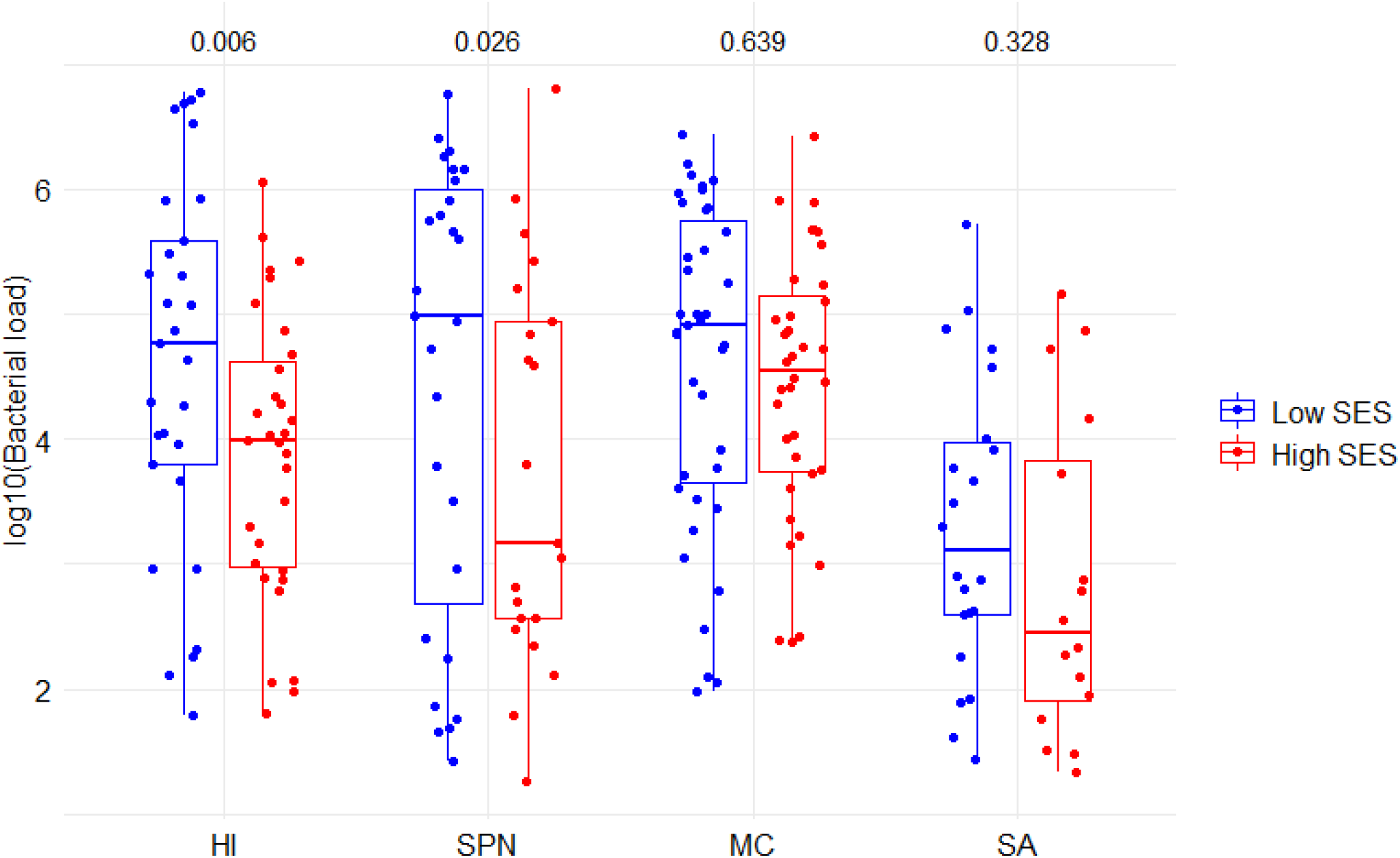
Bacterial loads in high and low SES school-aged children. All children for which bacterial colonization could be determined were included, thus n = 90 of which 46 low SES (blue) and 44 high SES (red) children. The boxes represent the interquartile range and the line within represents the median. The whiskers represent the 1.5 IQ of the upper and lower quartile bacterial load in each group. P-values are depicted above the corresponding boxplot and are derived with a multivariate regression model, correcting for age, sex and z-BMI. HI: *Hemophilus influenzae*, MC: *Moraxella catarrhalis*, SA: *Staphylococcus aureus*, SES: Socio-economic status, SPN: *Streptococcus pneumoniae*.

### Correlation between densities of the different bacteria

Multiple studies have shown that microbial species interact with each other and associations between different species have been described (20, 21). Correlation analysis showed that the H. influenzae density positively correlated with the M. catarrhalis density (β=0.53, *p*<0.001) and the bacterial load of S. pneumoniae negatively associated with S. aureus (β=-0.50, *p*=0.04) ***(Figure2)***. Furthermore, there was a trend for positive association between H. influenzae and S. pneumoniae density (β=0.34, *p*=0.07). No association was found between the other combinations of bacteria.

**Figure 2:**
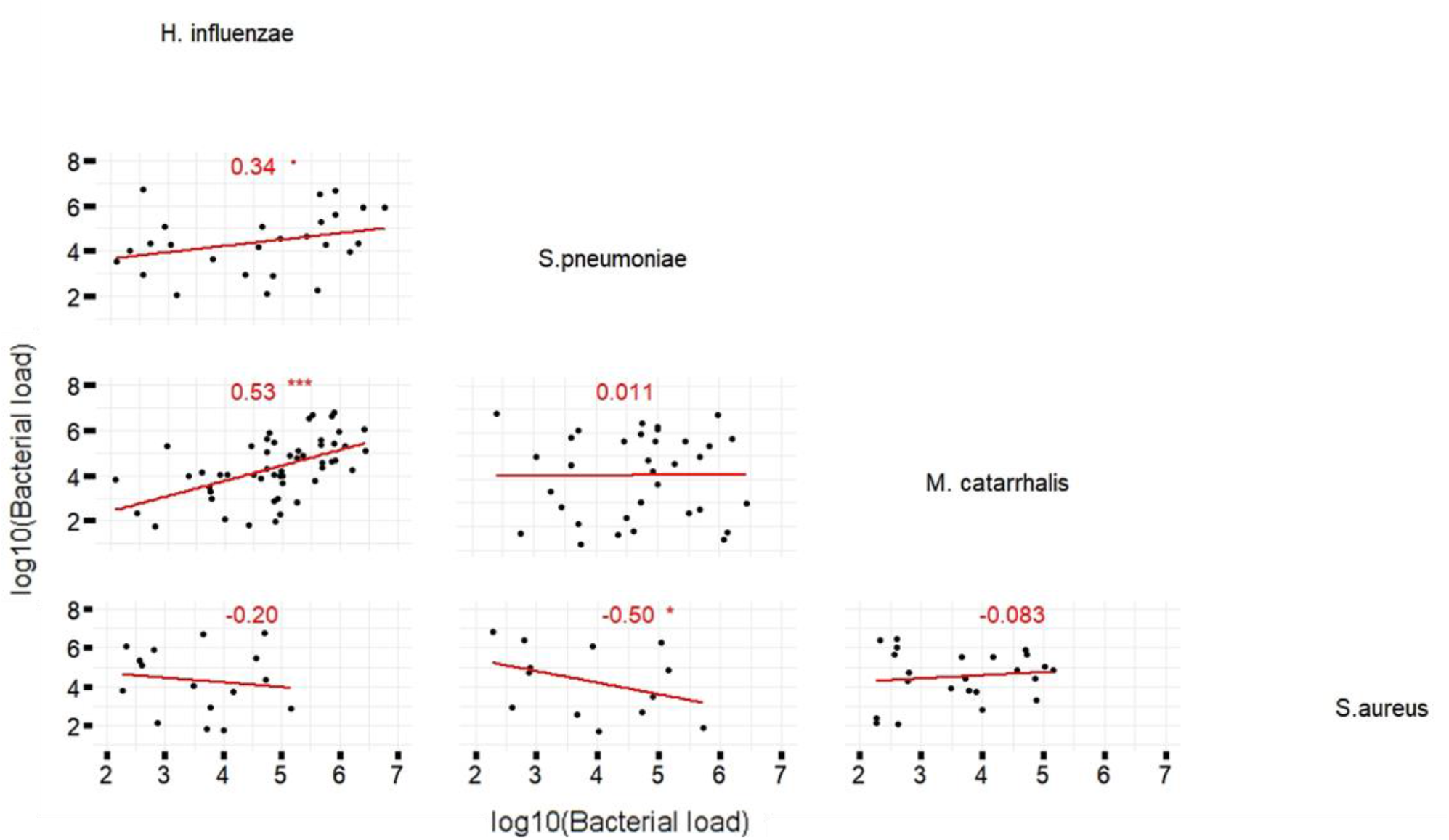
Correlation between the loads of different bacteria. All children for which bacterial colonization could be determined were included, thus n = 90. Left lower panel are the bivariate scatterplot of log_10_(bacterial loads) of the different bacteria with a regression line in red and for each plot the correlation (Pearson’s correlation) and as stars the significance level of this correlation ‘^***^’ for p-value < 0.001, ‘^**^’ for p-value ≤ 0.01, ‘^*^’ for p-value ≤ 0.05 and ‘^·^’ for p ≤ 0.10.

### IL-1beta concentrations are increased in the low SES compared to the high SES

To understand whether impaired mucosal responses might be responsible for the higher densities of colonized bacteria in low SES children, the concentration of 25 cytokines in nasal fluid were analyzed using a multiplex assay. Nine of these cytokines were detectable in the majority of samples: IL-18, IL-1alpha, IL-1beta, IL-1RA, IL-27, IL-4, IL-6, IL-7 and TNF-alpha. The other cytokines were below the limit of reliable detection in most samples ***(TableS3)***. Comparison of the cytokine levels between high and low SES showed that IL-1beta levels were significantly increased in the high SES, with a mean concentration of 338.8 in low and 102.3 in high SES after adjusting for confounders (β_*SES*_= − 0.518, p=0.048) ***(Figure3, TableS4)***.

**Figure 3:**
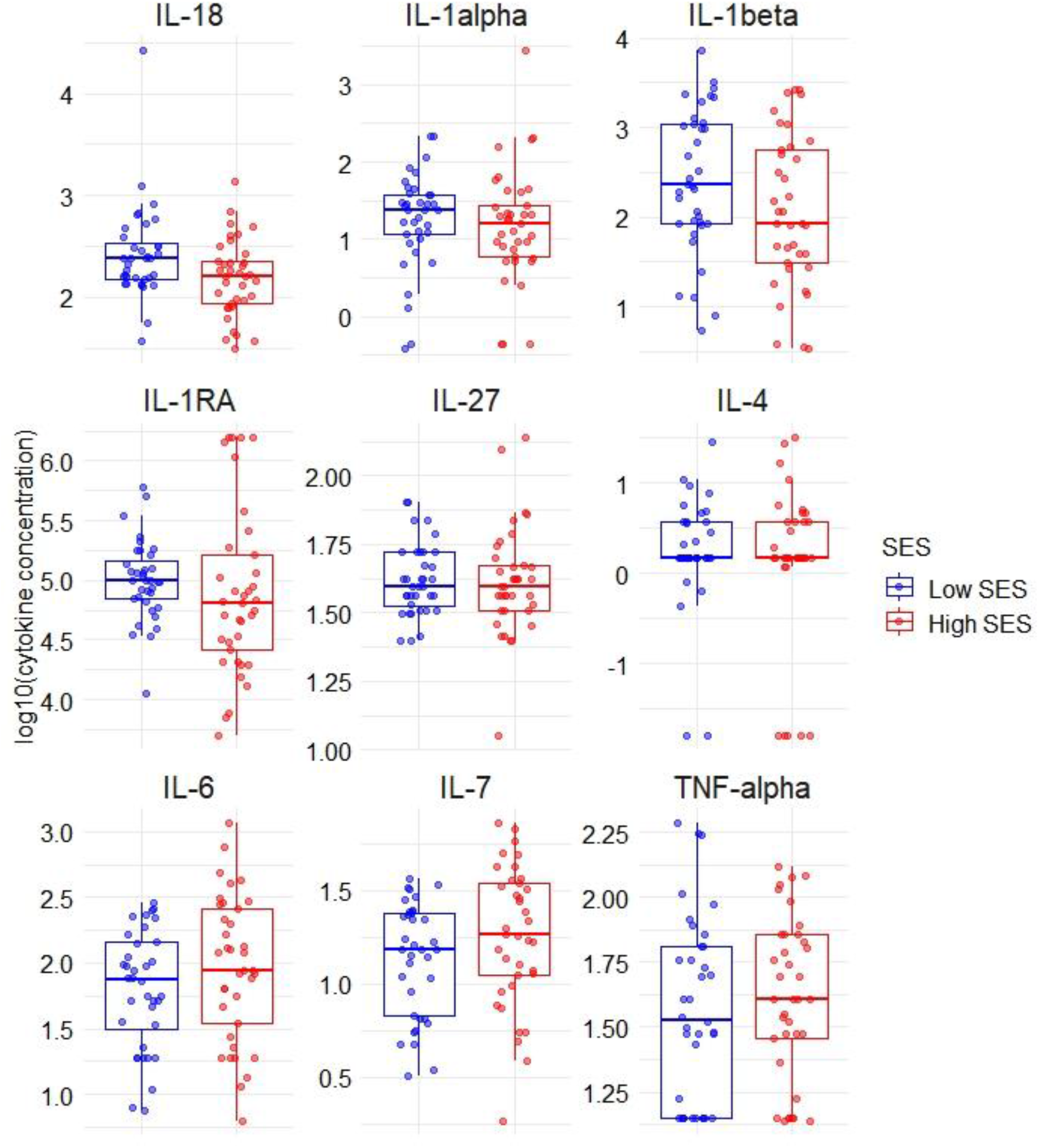
Nasal cytokine levels in high and low SES school-aged children. All children for which cytokines could be measured were included, thus n = 79 of which 39 low SES (blue) and 40 high SES (red) children. The boxes represent the interquartile range and the line within represents the median. Individuals are depicted by circles. The whiskers represent the 1.5 IQ of the upper and lower quartile. SES: Socio-economic status.

### IL-6 concentrations are increased in high SES children colonized by H. influenzae or S. pneumoniae

Similarly to PCA analysis, canonical correlation allows the integration of two multi-dimensional datasets to find common drivers of variation maximizing covariance between the datasets, allowing us to compare bacteria and cytokines on a global level (22). The densities of H. influenzae and S. pneumoniae and IL-1beta and IL-6 concentrations were identified as the main drivers of the correlation between bacteria and cytokines ***(TableS5)***. Indeed both H. influenzae and S. pneumoniae densities positively associated with IL-1beta and IL-6 levels ***(Figure4A-D)***. To quantify these correlations, a regression model including a priori confounders was used. The results showed that H. influenzae and S. pneumoniae densities are positively associated with IL-1beta (β_bacteria_ = 0.294, *p*=0.002, β_bacteria_=0.221, *p*=0.008, respectively) and IL-6 (β_bacteria_=0.190, *p*<0.001 and β_bacteria_=0.136, *p*=0.006) (TableS6). After adjusting for a priori confounders and the bacterial densities, IL-6 levels were increased in high compared to low SES in children colonized by H. influenzae (β_SES_=0.360, *p*=0.039) and S. pneumoniae (β_SES_=0.366, *p*=0.039), indicating that IL-6 levels were increased in high compared to low SES at any given density of these bacteria. To examine whether SES affected the strength and direction of this association, a regression model including the interaction between these bacterial density and cytokine levels was performed, which showed that this was not significant ***(TableS7)***. Thus, children from both high and low SES had dose-dependent cytokine responses to H. influenzae or S. pneumoniae colonization, but these responses were stronger in high SES children for IL-6.

**Figure 4:**
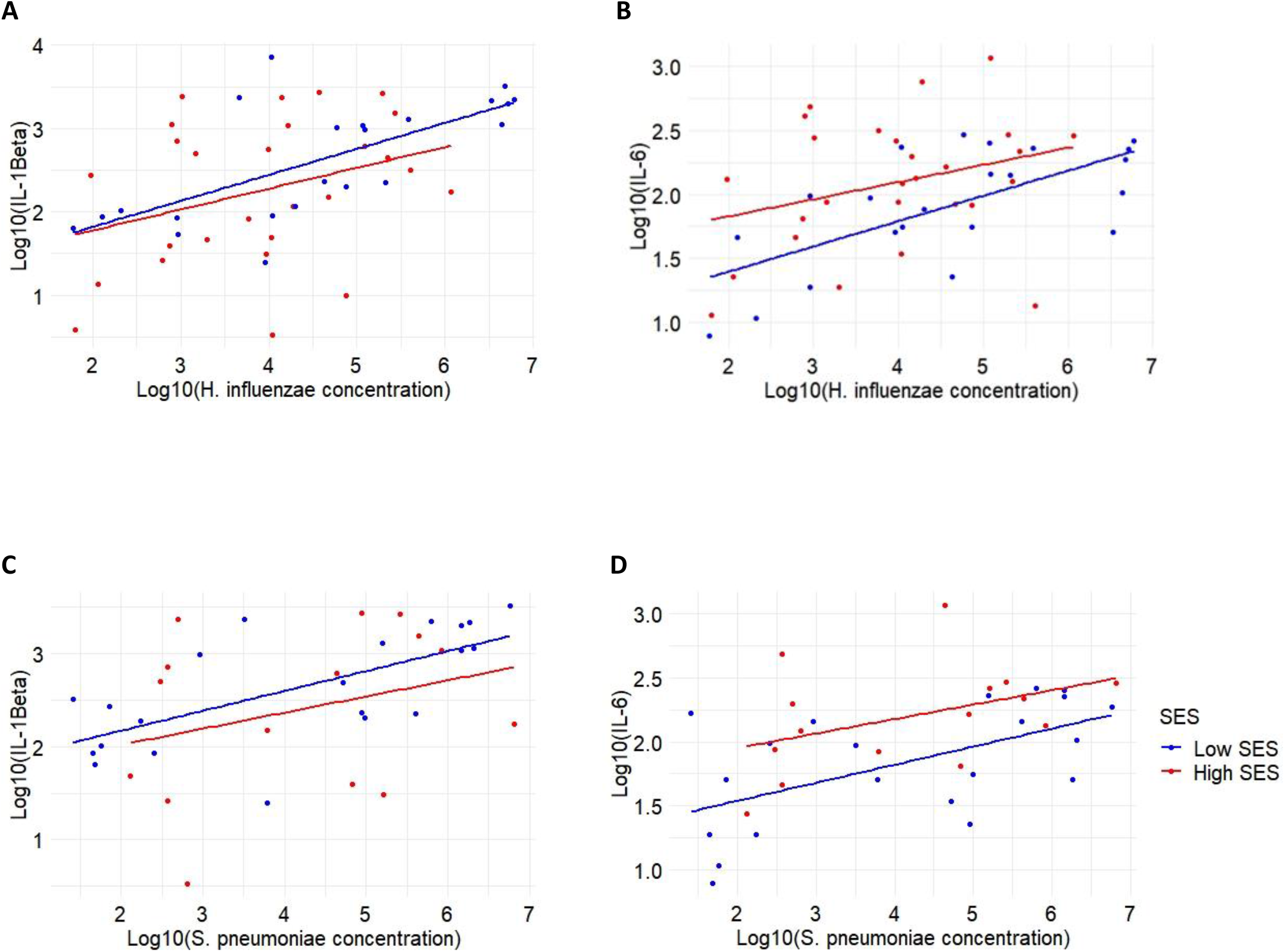
Association between *H. influenzae* (A and B) or *S. pneumoniae* concentration (C and D) and IL-1beta and IL-6 concentration in high and low SES school-aged children. For all models with *H. influenzae* only children colonized are included thus n = 49, which includes 27 high SES children and 22 low SES children. For all models with *S. pneumoniae* only children colonized are included thus n = 36, which includes 15 high SES children and 21 low SES children.

### IL-1RA and IL-1beta response to S. aureus carriage differs between high and low SES

Since a limited number of children were colonized with S. aureus and the presence and density was not associated with that of the other bacteria, responses to colonization by S. aureus were not likely to be identified by the canonical correlation analysis. Therefore, the relation between S. aureus carriage status and cytokine concentrations was further examined, whereby IL-1 cytokines were of main interest since these cytokines were shown previously to be important for S. aureus control (8). In the low SES, the concentration of IL-1RA seemed to be higher in in S. aureus carriers compared to non-carriers, whereas in high SES IL-1RA levels were lower in the carriers compared to non-carriers ***(Figure5A)***. A similar pattern was observed for IL-1beta ***(Figure5B)***. To quantify these observations, we performed a regression analysis including the a priori confounders, showing a significant difference between high and low SES for IL-1RA (β_interaction_=-0.700, *p*=0.007), and a similar trend for IL-1beta (β_interaction_=-0.700, *p*=0.059) ***(TableS8)***. Since IL-1beta and IL-1RA compete for the same receptor and the activity of IL-1beta is affected by the levels of IL-1RA, the ratio between IL-1RA and IL-1beta was determined ***(Figure5C)***. The interaction between S. aureus carriage and SES was not seen for the ratio between these cytokines (β_interaction_=0.074, *p*=0.859), but the IL-1RA/IL-1beta ratio tended to be higher in high compared to low SES (β_SES_=0.465, *p*=0.078) ***(TableS8)***. The pattern observed for IL-1beta and IL-1RA did not exist for other IL-1 cytokines, including IL-1alpha ***(FigureS1)***.

**Figure 5:**
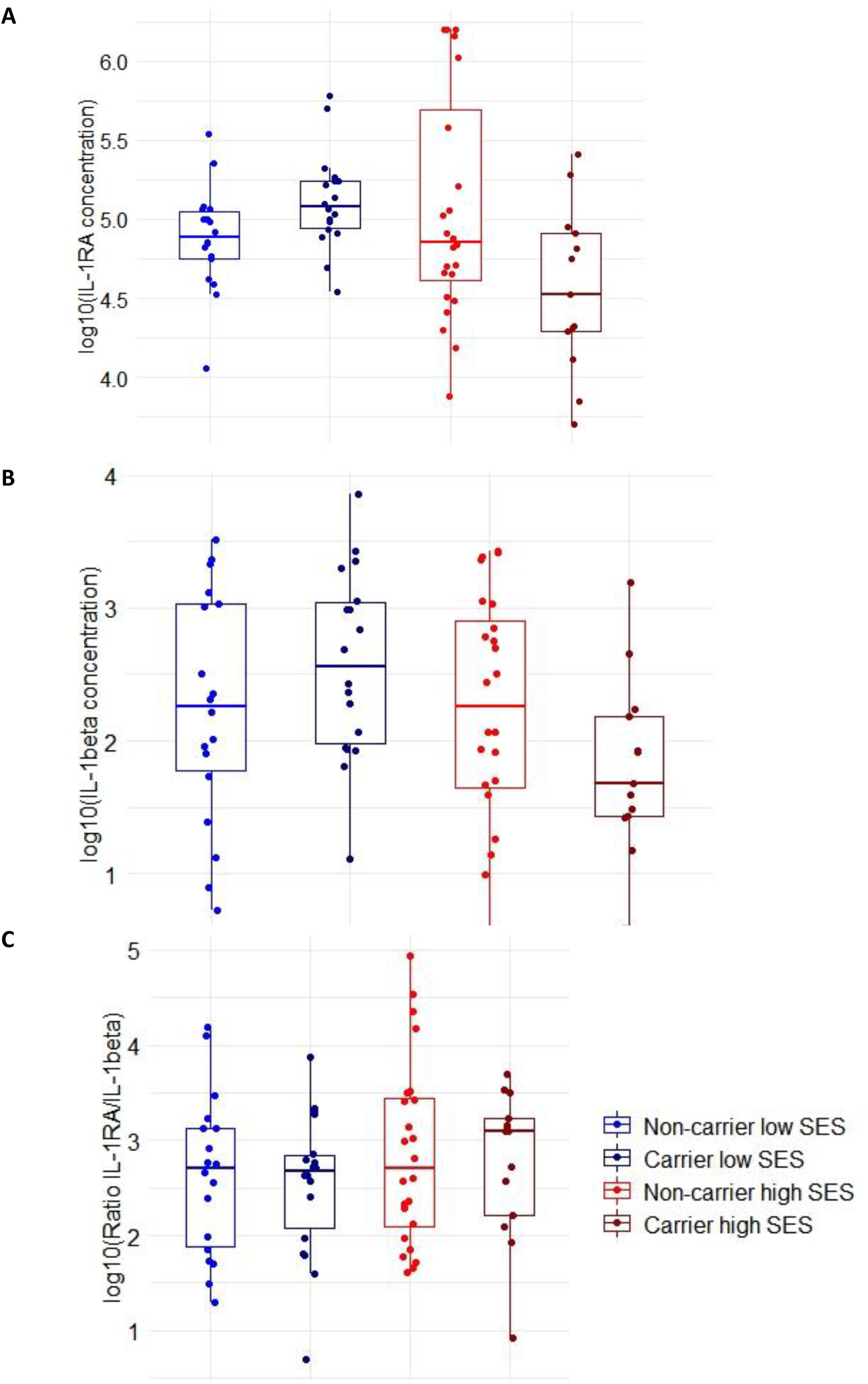
The IL-1RA and IL-1Beta concentrations *in S. Aureus* carrier and non-carrier in high and low SES. All children for which both cytokines and bacterial colonization could be measured were included, thus n = 73 of which 36 low SES and 37 high SES children. The boxes represent the interquartile range and the line within represents the median. The whiskers represent the 1.5 IQ of the upper and lower quartile concentration of the cytokine. SES: Socio-economic status.

## Discussion

This is the first study evaluating the nasal cytokine response in relation to bacterial colonization in high and low SES children. This study showed increased densities of H. influenzae and S. pneumoniae in low compared to high SES. The densities of these bacteria were positively associated with IL-1beta and IL-6 levels. After correcting for bacterial density, IL-6 levels were increased in high SES, indicating that IL-6 levels were higher at any given density of H. influenzae and S. pneumoniae in high compared to low SES. Furthermore, increased levels of IL-1RA were found in S. aureus carriers compared to non-carriers in low SES, whereas in high SES IL-1RA were higher in the non-carriers. A similar pattern was observed for IL-1beta but not for other cytokines. The IL-1RA/IL-1beta ratio was not affected by S. aureus carrier status, but tended to be increased in high compared to low SES.

SES has been identified as risk factor for increased nasopharyngeal carriage in a number of studies (4, 11-14). A previous study in Indonesia found increased densities of S. aureus in children living in semi-rural compared to urban areas. Furthermore, they identified family income as a risk factor for pneumococcal carriage densities and showed that low maternal education was associated with increased H. influenzae densities (14). The results of the current study are in line with these previous findings, however in contrast to the study Fadlyana et al. we included children from different SES within one urban center, thereby providing unique insights into the role of SES on nasopharyngeal bacterial carriage and densities in children.

Without taking bacterial colonization status into account, only IL-1beta was significantly different between high and low SES, while there was a large dynamic range in cytokine levels observed, indicating that local factors are important for driving cytokine levels at the mucosa. Indeed, densities of H. influenzae and S. pneumoniae were positively associated with IL-1beta and IL-6 levels, highlighting the importance of the activation of the IL-1 signaling pathway in response to bacterial colonization. The importance of the IL-1 cytokine signaling in the clearance of S. pneumoniae has been shown in mice and reduced IL-1 responses have been suggested to be permissive for persistent colonization during infancy (10, 23). Moreover, high levels of IL-6 and IL-1beta were found in response to Non-typeable H. influenzae in individuals suffering from chronic suppurative lung disease (24).

After adjusting for bacterial densities, increased IL-6 levels were observed in children colonized by H. influenzae or S. pneumoniae from high compared low SES, whereas this was not observed for IL-1beta. This might be explained by the role of IL-6 in the IL-1 signaling pathway and its capacity to drive promote the differentiation of Th17-cells. Whereas IL-1beta is primarily initiates the IL-1 signaling, IL-6 production is one of the numerous downstream effects of the IL-1 signaling pathway. Furthermore, high levels of IL-6 are known to promote differentiation of CD4^+^-cells to form Th17-cells, a cell type that has been shown essential for the clearance of S. pneumoniae in mice (25). A study analyzing the Th17-cells and cytokines in adenoidal tissue from S. pneumoniae-positive and S. pneumoniae-negative children found an increased number of Th17-cells and higher levels of IL-17A and IL-6 in S. pneumoniae-negative compared to S. pneumoniae-positive children (26). In the current study increased IL-6 levels were observed in the high SES, whereas the density of H. influenzae and S. pneumoniae was reduced, supporting the role of IL-6 in the control of these bacteria. The IL-17A levels were below the level of detection in most samples collected in this study, however very low levels of IL-17A might be enough to activate local T-cells and therefore be involved in the bacterial clearance.

In the current study in the low SES increased IL-1beta and IL-1RA levels were observed in S. aureus carriers compared to non-carriers, whereas in the high SES decreased levels were found in carriers. The ratio between IL-1RA and IL-1beta was not affected by the carrier status but tended to be decreased in low compared to high SES after adjusting for S. aureus carriage. A human nasal inoculation with S. aureus showed that IL-1beta was upregulated after inoculation in individuals able to clear the bacteria compared to individuals, but found no difference in IL-1RA levels. Furthermore, they showed that IL-1RA/IL-1beta ratio was significantly decreased in individuals with persistent S. aureus and proposed this ratio as metric for the clearance of S. aureus in the nasal mucosa rather than the expression of individual cytokines (8). In the current study, the IL-1RA/IL-1beta ratio tended to be increased in high compared to low SES, indicating that the response to S. aureus in low SES children might be reduced. No significant differences in the IL-1RA/IL-1beta ratio between carriers and non-carriers were found, however, it should be kept in mind that the duration of the colonization was not measured in the current study and some of the carriers would be able to clear S. aureus over time. To elucidate the relation between SES and S. aureus persistence, longitudinal studies are thus needed.

A factor that can affect the bacterial colonization and cytokine responses and which can be impacted by SES is the vaccination status against pneumococcal and H. influenzae type B (Hib) diseases. The children in our study were not vaccinated through their national childhood vaccination program. However, in a private clinic on payment additional vaccines can be administered, which predominantly involves the Hib vaccine. Although vaccination status might differ between SES its effect on the bacterial load detected here is probably limited, since the (sero)types prevalent in Indonesia only partially match with the available vaccines. A cross-sectional study amongst 302 young children in Indonesia in 2016, thus after Hib vaccine was incorporated in the national program but before the pneumococcal conjugate vaccine (PCV13) was introduced, showed a carriage rate of 27.5% of H. influenzae, but none of these isolates was type B (27). Moreover, before the introduction of Hib vaccination it was shown that Hib vaccination would not have an effect on the pneumonia incidence in Indonesia whereas it in Africa and South America significantly did, indicating that in Indonesia other respiratory pathogens are responsible for LRTIs (28). Moreover, study by Dunne et al. showed that only 46% of the pneumococcal isolates obtained from Indonesian children are covered by the PCV13 (27). While location-specific serotype circulation is of course possible, one can expect that the isolates found in our study are also (sero)types that will only partly be covered by the vaccine, which limits the effect of differences in vaccination status between high and low SES on the results.

One of the limitations of the current study is that only four pathogenic bacteria were measured in a cross-sectional manner, whereas it is known that viral coinfections and microbiota in general are also important in shaping responses. Moreover, future studies should assess changes and responses longitudinally. In addition, while school-going children are important drivers of transmission within communities, most childhood pneumonia occurs before the age of 5, therefore it would also be of interest to study mucosal immune responses in infants in future studies.

To conclude, the results in the current study indicate that the local immune response to nasopharyngeal bacterial colonization is altered by SES. Since nasopharyngeal carriage of these bacteria precedes disease and local immune responses are important in controlling potential pathogenic bacteria elucidating the relation between the bacterial colonization and local cytokine response is an important first step in understanding the bacteria-host relationship. Insights in this relationship are important for the development of vaccine strategies and treatment options and will eventually lead to reduced LRTIs worldwide.

## Data Availability

All data produced in the present work are contained in the manuscript

## Presentation of the work

This work has not been previously presented.

## Acknowledgements

The authors would like to thank Koen Stam and Mikhael Manurung for their advice on the statistical analysis and all participants involved in this study as well as all students of the LUMC and Hasanuddin University for their efforts on sample collection in participating primary schools in Makassar. We thank Hasanuddin University Medical Research Center (HUM-RC) for laboratory facilities during sample collections, and Hasanuddin University for their support.

## Author contributions

Study design: SJ, MY, ES

Local organization of the study: SW, AA, MM, ES

Sample collection: MD, AA, MM, SJ, SW

Experimental measurements: MD, SA

Data analysis: MD, SA, SJ

Data interpretation: MD, SA, SJ

Writing manuscript: MD, SJ, MY

## Figure legends

**Supplementary figure S1: IL-1α concentration in *S. aureus* carrier and non-carrier in high and low SES.** All children for which both cytokines and bacterial colonization could be measured were included, thus n = 73 of which 36 low SES and 37 high SES children. SES: Socio-economic status.

