## Supplemental table 1 for "Differences in bacterial colonization and mucosal responses between high and low SES children in Indonesia"

**Table S1: Characteristics of sample population**

|  | |  | | Low SES  (n= 50) | High SES (n=48) | p-value |
| --- | --- | --- | --- | --- | --- | --- |
| Age (in years, mean, SD) | |  | | 8.34 ± 1.40 | 7.85 ± 1.17 | 0.061 |
| Sex (female %, n/N) | |  | | 50.0 (25/ 50) | 45.8 (22/48) | 0.680 |
| z-BMI (mean, SD) | |  | | -1.74 ± 1.51 | 0.44 ± 1.82 | <0.001 |
| Education father (high %, n/N) | |  | | 8.3 (4/48) | 84.4 (38/45) | <0.001 |
| Maternal education (high %, n/N) | | |  | 8.3 (4/48) | 73.9 (34/46) | <0.001 |
| Floor material (ceramics %, n /N) | | |  | 36.7 (18/49) | 95.7 (44/46) | <0.001 |
| Wall material (brick or concrete %, n/N) | |  | | 53.1 (26/49) | 100 (46/46) | <0.001 |
| Toilet (private inside %, n/N) | |  | | 83.3 (40/48) | 100 (46/46) | 0.004 |
| Helminth infection by microscope (%, n/N) | |  | | 6.0 (3/43) | 0.0 (0/33) | 0.122 |
| Bacterial colonization (carrier % , n/N) | |  | |  |  |  |
|  | *H. Influenzae* | | | 63.0 (29/46) | 70.5 (31/44) | 0.456 |
|  | *S. Pneumoniae* | | | 56.5 (26/46) | 47.7 (21/44) | 0.404 |
|  | *M. Catarrhalis* | | | 80.4 (37/46) | 81.8 (36/44) | 0.867 |
|  | *S. Aureus* | | | 47.8 (22/46) | 36.4 (16/44) | 0.271 |
| Number of bacteria (%, n/N) | | | |  |  |  |
|  | No bacteria | | | 0.0 (0/46) | 2.3 (1/44)* | 0.865 |
|  | One bacterium | | | 21.7 ( 10/46) | 20.5 (9/44) |  |
|  | Two bacteria | | | 28.3 (13/46) | 31.8 (14/44) |  |
|  | Three bacteria | | | 30.4 (14/46) | 29.5 (13/44) |  |
|  | All four bacteria | | | 19.6 (9/46) | 15.9 (7/44) |  |

The number of positives (n) of the total number (N). SD: standard deviation. SES: socio-economic status. CI: confidence interval. Student’s t-test for continuous variables and Pearson’s Chi-square for binary/categorical variables. *qPCR with 16S was performed for this child to confirm DNA extraction was successful
