## Supplemental table 3 for "Differences in bacterial colonization and mucosal responses between high and low SES children in Indonesia"

| Cytokine | Number of samples in range (%)^1^ | Number of samples in range in high SES  (%)^1^ | Number of samples in range in low SES  (%)^1^ | Number of samples in reliable range (%) ^2^ | Number of samples in reliable range in high SES (%) ^2^ | Number of samples in reliable range in low SES (%) ^2^ |
| --- | --- | --- | --- | --- | --- | --- |
| GM-CSF | 28 (35%) | 17 (43%) | 11 (28%) | 3 (4%) | 3 (8%) | 0 |
| IFN-alpha | 7 (9%) | 1 (3%) | 6 (15%) | 0 | 0 | 0 |
| IFN-gamma | 50 (63%) | 27 (68%) | 23 (59%) | 13 (16%) | 8 (20%) | 5 (13%) |
| IL-2 | 17 (22%) | 13 (33%) | 4 (10%) | 7 (9%) | 6 (15%) | 1 (3%) |
| IL-5 | 17 (22%) | 8 (20%) | 9 (23%) | 4 (5%) | 3 (8%) | 1 (3%) |
| IL-9 | 51 (65%) | 26 (65%) | 25 (64%) | 4 (5%) | 4 (10%) | 0 |
| IL-10 | 45 (57%) | 25 (63%) | 20 (51%) | 7 (9%) | 6 (15%) | 1 (3%) |
| IL-12p70 | 49 (62%) | 26 (65%) | 23 (59%) | 4 (5%) | 3 (8%) | 1 (3%) |
| IL-13 | 35 (44%) | 16 (40%) | 19 (49%) | 13 (16%) | 8 (20%) | 5 (13%) |
| IL-15 | 32 (41%) | 18 (45%) | 14 (36%) | 4 (5%) | 4 (10%) | 0 |
| IL-17A | 56 (71%) | 29 (73%) | 27 (69%) | 7 (9%) | 6 (15%) | 1 (3%) |
| IL-21 | 44 (56%) | 20 (50%) | 24 (62%) | 8 (10%) | 5 (13%) | 3 (8%) |
| IL-22 | 27 (34%) | 12 (30%) | 15 (38%) | 10 (13%) | 5 (13%) | 5 (13%) |
| IL-23 | 7 (9%) | 6 (15%) | 1 (3%) | 0 | 0 | 0 |
| IL-31 | 5 (6%) | 3 (8%) | 2 (5%) | 0 | 0 | 0 |
| TNF-beta | 7 (9%) | 4 (10%) | 3 (8%) | 0 | 0 | 0 |

**Table S3: Number of samples in range and in reliable range per cytokine**

All children for which cytokines could be measured were included, thus n = 79 of which 39 low SES and 40 high SES children.

^1^ Number of samples for which the samples could be determined based on their position in relation to the standard curve or that were out of range but could be estimated based on the MFI value.

^2^ Reliable range (often) corresponds with above the seventh standard and is determined based on the standard curve for each cytokine. None of the cytokines had levels that were out of range above the standards
