## Supplemental table 4 for "Differences in bacterial colonization and mucosal responses between high and low SES children in Indonesia"

**Table S4: Results of regression models cytokine levels adjusted for *a priori* confounders**

|  | | | Mean cytokine concentration | | | Coefficient SES (95% CI) | P-value | |
| --- | --- | --- | --- | --- | --- | --- | --- | --- |
| IL-18 | | | |  |  |  | | |
|  | | Low SES | 741.3 | | | Reference | |  |
|  | | High SES | 468.8 | | | -0.199 ( -0.442 ; 0.0439) | | 0.113 |
| IL-1alpha | | | |  |  |  | | |
|  | | Low SES | 9.984 | | | Reference | | 0.389 |
|  | | High SES | 6.180 | | | -0.186 ( -0.605 ; 0.234) | |  |
| IL-1beta | | | |  |  |  | | |
|  | | Low SES | 338.8 | | | Reference | |  |
|  | | High SES | 102.3 | | | -0.518 ( -1.02 ; -0.0149) | | **0.048** |
| IL-1RA | | | |  |  |  | | |
|  | Low SES | | 75,857 | | | Reference | |  |
|  | High SES | | 67,608 | | | -0.0494 ( -0.374; 0.275) | | 0.766 |
| IL-27 | | | |  |  |  | | |
|  | Low SES | | 42.66 | | | Reference | |  |
|  | High SES | | 49.32 | | | 0.0626 (-0.0360; 0.161) | | 0.218 |
| IL-4 | | | |  |  |  | | |
|  | Low SES | | 2.455 | | | Reference | |  |
|  | High SES | | 2.786 | | | 0.0553 (-0.377; 0.488) | | 0.803 |
| IL-6 | | | |  |  |  | | |
|  | Low SES | | 85.11 | | | Reference | |  |
|  | High SES | | 116.95 | | | 0.138 (-0.164; 0.439) | | 0.374 |
| IL-7 | | | |  |  |  | | |
|  | Low SES | | 16.60 | | | Reference | |  |
|  | High SES | | 21.18 | | | 0.106421 (-0.0852; 0.298) | | 0.280 |
| TNF-alpha | | | |  |  |  | | |
|  | Low SES | | 53.70 | | | Reference | |  |
|  | High SES | | 64.41 | | | 0.0788 (-0.119; 0.276) | | 0.437 |

All children for which cytokines could be measured were included, thus n = 79 of which 39 low SES and 40 high SES children. Mean cytokine concentration is the geometric mean in high and low SES after adjusted for *a priori* confounders including age (in years), sex and z-BMI.
