## Supplemental table 5 for "Differences in bacterial colonization and mucosal responses between high and low SES children in Indonesia"

**Table S5: Canonical correlation analysis bacterial loads**

|  | |  | Standardized coefficients |
| --- | --- | --- | --- |
| Bacterial load | |  |  |
|  | *H. influenzae* | | **-0.877** |
|  | *S. pneumoniae* | | **-0.240** |
|  | *M. catarrhalis* | | 0.109 |
|  | *S. aureus* | | -0.016 |
| Cytokine concentration | |  |  |
|  | IL-1alpha | | 0.097 |
|  | IL-1beta | | **-0.791** |
|  | IL-1RA | | 0.037 |
|  | IL-4 | | 0.080 |
|  | IL-6 | | **-0.311** |
|  | IL-7 | | -0.028 |
|  | IL-18 | | 0.309 |
|  | IL-27 | | 0.173 |
|  | TNF-alpha | | -0.141 |

All children for which both cytokines and bacterial colonization could be measured were included, thus n = 73 of which 36 low SES and 37 high SES children. Standardized coefficient of the canonical correlation analysis first dimension with *p*=0.042 using Wilks’ Lambda. In bold the variables that predominantly drive the correlation and that will be used for further analysis. Both bacterial loads and cytokine concentrations are log_10_ transformed.
