## Supplemental table 6 for "Differences in bacterial colonization and mucosal responses between high and low SES children in Indonesia"

|  | |  | Intercept  (95% CI) | Coefficient bacterial load (95% CI) | P-value  bacterial load | Coefficient SES  (95% CI) | P-value SES |
| --- | --- | --- | --- | --- | --- | --- | --- |
| *H. influenzae* | |  |  |  |  |  |  |
|  | IL-1beta | | 1.12 (-0.661 – 2.91) | 0.294 (0.119 – 0.470) | 0.002 | -0.123 (-0.742 – 0.496) | 0.699 |
|  | IL-6 | | 1.14 (0.189 – 2.09) | 0.190 (0.096 – 0.284) | <0.001 | 0.360 (0.029 – 0.284) | 0.039 |
| *S. pneumoniae* | |  |  |  |  |  |  |
|  | IL-1beta | | 1.48 (-0.413 – 3.41) | 0.221 (0.069 – 0.373) | 0.008 | -0.154 (-0.720 – 0.411) | 0.596 |
|  | IL-6 | | 1.14 (0.032 – 2.27) | 0.136 (0.047 – 0.225) | 0.006 | 0.366 (0.03 – 0.697) | 0.039 |

**Table S6: Relation between *H. influenzae* or *S. pneumoniae* bacterial load and the IL-1beta or IL-6 concentrations**

Adjusted for *a priori* confounders age (in years), sex and z-BMI. Both bacterial loads and cytokine concentrations are log10 transformed. For all models with *H. influenzae* only children colonized are included thus n = 49, which includes 27 high SES children and 22 low SES children. For all models with *S. pneumoniae* only children colonized are included thus n = 36, which includes 15 high SES children and 21 low SES children**.**
