## Supplemental table 7 for "Differences in bacterial colonization and mucosal responses between high and low SES children in Indonesia"

**Table S7: Regression model *H. influenzae* and *S. pneumoniae* load and the IL-1beta or IL-6 concentrations with interaction term**

|  | |  | | Intercept (95% CI) | Coefficient bacterial load (95% CI) | P-value bacterial load | Coefficient SES (95% CI) | P-value SES | Coefficient interaction (95% CI) | p-value interaction |
| --- | --- | --- | --- | --- | --- | --- | --- | --- | --- | --- |
| *H. influenzae* | |  | |  |  |  |  |  |  |  |
|  | IL-1beta | | | 1.01 (-0.951 – 2.96) | 0.316 (0.089 – 0.542) | 0.009 | 0.105 (-1.58 – 1.71) | 0.899 | -0.05 (-0.393 – 0.288) | 0.764 |
|  | IL-6 | | | 1.12 (0.076 – 2.17) | 0.194 (0.073 – 0.314) | 0.003 | 0.398 (-0.461 – 1.26) | 0.369 | -0.01 (-0.191 – 0.173) | 0.924 |
| *S. pneumoniae* | | |  |  |  |  |  |  |  |  |
|  | IL-1beta | | | 1.46 (-0.554 – 3.47) | 0.228 (0.05 – 0.402) | 0.012 | -0.056 (-1.40 – 1.29) | 0.935 | -0.025 (-0.330 – 0.280) | 0.875 |
|  | IL-6 | | | 1.10 (-0.076 – 2.28) | 0.144 (0.042 – 0.246) | 0.010 | 0.486 (-0.300 – 1.27) | 0.236 | -0.030 (-0.209 – 0.148) | 0.743 |

Adjusted for *a priori* confounders age (in years), sex and z-BMI. Both bacterial loads and cytokine concentrations are log10 transformed. For all models with H. influenzae only children colonized are included thus n = 49, which includes 27 high SES children and 22 low SES children. For all models with S. pneumonia only children colonized are included thus n = 36, which includes 15 high SES children and 21 low SES children**.**
