## Supplemental table 8 for "Differences in bacterial colonization and mucosal responses between high and low SES children in Indonesia"

|  | | Intercept  (95% CI) | | Coefficient carrier status (95% CI) | P-value carrier status | Coefficient SES  (95% CI) | P-value SES | Coefficient interaction (95% CI) | p-value interaction |
| --- | --- | --- | --- | --- | --- | --- | --- | --- | --- |
|  | IL-1RA | 4.59 (3.75 – 4.44) | 0.199 (-0.144 – 0.543) | | 0.260 | 0.228 (-0.150 – 0.605) | 0.242 | -0.700 (-1.19 – -0.207) | **0.007** |
|  | IL-1beta | 2.21 (0.851 – 3.56) | 0.251 (-0.300 – 0.801) | | 0.376 | -0.206 (-0.811- 0.340) | 0.508 | -0.772 (-1.56 – 0.016) | 0.059 |
|  | Ratio IL-1RA/ IL-1beta * | 2.39 (1.00 – 3.78) | -0.051 (-0.615 - 0.513) | | 0.860 | 0.433 (-0.187 – 1.053) | 0.175 | 0.074 (-0.733 – 0.881) | 0.859 |
|  | Ratio IL-1RA/ IL-1beta ** | 2.36 (1.02 – 3.69) | -0.017 (-0.432 – 0.399) | | 0.937 | 0.465 (-0.044 – 0.975) | 0.078 |  |  |

**Table S6: Relation between *S. Aureus* carrier status and the IL-1beta or IL-1RA concentrations with interaction term**

Adjusted for *a priori* confounders age (in years), sex and z-BMI. Cytokine concentrations are log10 transformed. All children for which both cytokines and bacterial colonization could be measured were included, thus n = 73 of which 36 low SES and 37 high SES children. ** log10 transformed and with interaction term, * log10 transformed without interaction term
